# Direct-to-Consumer Chat-Based Remote Care Before and During the COVID-19 Outbreak

**DOI:** 10.1101/2020.07.14.20153775

**Authors:** Dan Zeltzer, Alina Vodonos, Edo Paz, Roy Malka

## Abstract

**Objective:** To compare the patient population, common complaints, and physician recommendations in direct-to-consumer chat-based consults, before and during the COVID-19 outbreak.

**Data sources:** Data on patient characteristics, patient complaints, and physician recommendations from 36,864 chat-based telemedicine consults with physicians in an online-clinic by patients from across the United States between April 2019 and April 2020.

**Study Design:** We perform a retrospective analysis comparing patient characteristics, visit characteristics, and physician recommendation before and after the COVID-19 outbreak. We examine patient age and gender, visit time, patient chief complains, and physician medical recommendation (including prescription drugs, reassurance, and referrals).

**Principal Findings:** Before March 2020, most patients were female (75 percent) and 18–44 years old (89 percent). Common complaints such as abdominal pain, dysuria, or sore throat suggested minor acute conditions. Most cases (67 percent) were resolved remotely, mainly via prescriptions; a minority were referred. Since March 2020, the COVID-19 emergency has led to a sharp (fourfold) increase in case volume, including more males (from 25 to 29 percent), patients aged 45 and older (from 11 to 17 percent), and more cases involving mental health complaints and complaints related to COVID-19. Across all symptoms, significantly more cases (78 percent) have been resolved remotely.

**Conclusions:** The COVID-19 outbreak in the United States has been associated with a sharp increase in the use of chat-based telemedicine services, including by new patient demographics, an increase in both COVID-19 and mental health complains, and an increase in remote case resolutions.

## Introduction

In response to the need to maintain social distance, the Coronavirus Diseases 2019 (COVID-19) crisis has catalyzed a rapid expansion in telemedicine services, which provide the opportunity to maintain access to and continuity of medical care while reducing the potential for contagion. In March, the United States federal government, the United Kingdom government, and many other healthcare systems throughout the world substantially expanded the provision and coverage of care via telemedicine, leading to a surge in both supply and demand for it.^1,2^ The growth of telemedicine enabled and accelerated the use of different models of remote care, raising the question of what will become of these new approaches to treatment once the immediate COVID-19 crisis has passed.^3^

A care model that was popular even before the current crisis involves chat-based direct-to-consumer interactions based on mobile technology.^3,4^ Mobile apps and patient portals allow instantaneous remote written consults with providers, synchronously and asynchronously.^5,6^ Consults are typically initiated by patients and include symptom intake followed by an interaction with a provider, usually for acute conditions. The benefits of chat-based consults include convenience and ease of access relative not only to office-based care by eliminating travel but also to remote care via video and voice conversations, as these written consults can be used privately even in crowded spaces, such as during a commute. Services include both asynchronous and synchronous encounters with providers that are sometimes augmented by algorithms that facilitate symptom intake or diagnostics. Patient-initiated remote care can be seen as the digital evolution of on-demand walk-in care that has been expanding in recent years in various settings such as retail clinics and urgent care centers, that resolve minor acute conditions and triage for more urgent or complex cases.

This study analyzes the use of direct-to-consumer, chat-based care in one online clinic that has been serving patients in more than 40 US states since April 2019. We study both usage before the COVID-19 crisis and changes brought about by the crisis through the emergency lockdowns that restricted access to traditional care settings. Understanding past and current use of chat-based remote care can inform policy discussions about potential roles for this care technology during current continuing efforts to mitigate COVID-19 and in the future once this crisis has passed.

## Setting and Data

We study an online clinic affiliated with K Health, a technology startup, that has been offering chat-based synchronous consults since April 2019.^7^ Consults can be initiated by patients from anywhere in the United States through a mobile app available through app stores. Upon log in, patients report their age, gender, and current symptoms through an automated and adaptive structured questionnaire generated by a machine learning algorithm. After completing a conversation with the app, users are provided with information regarding the potential conditions they may have, which is automatically generated by an algorithm. At that point, patients can request to chat with a doctor, for an out-of-pocket payment (during the study period, the fee was up to $20 per visit). Requests are routed to the first available physician in a US-based online clinic operated by K Health.

The assigned physician reviews a summary of the patient intake and further interacts with the patient via free text-based chat. Physicians can provide reassurance and self-care instructions, recommend over-the-counter drugs, prescribe drugs electronically, and order diagnostic labs. Physicians may also refer patients to an office-based primary care physician or specialist or, if they deem it appropriate, to an urgent care center or an emergency department.

## Study Design and Methods

### Study Sample and Key Variables

We perform a retrospective analysis of electronic health records of chat-based telemedicine consults that were completed through the K Health mobile app from April 2019 through April 2020. Our sample consists of 36,864 completed chat consults by patients aged 18–65. We exclude a small number of cases that were discontinued before a recommendation had been made due to technical problems. Data on each visit include the patient age, gender, the chief complaint reported by the patient during the intake, the time the patient initiated the chat (in the patient’s local time), and the physician recommendation at the end of the chat. Based on the physician recommendation, we consider cases as “resolved,” if they were concluded with a physician reassurance, prescription, or a lab order. We consider cases as “referred” if the physician referred the patient to a primary care physician, other physician specialists, urgent care, or an emergency department.

### Study Period

Our study period is from April 2019 through April 2020. Therefore, our sample includes consults that occurred both before and during the COVID-19 crisis, and thus allow us to compare patient and consult characteristics between these periods. While the first COVID-19 case was identified in the United States in late January 2020, it was not until March that most activities, including primary care, were affected in most states. As seen in Exhibit 1, the surge in the weekly number of consults started only after a national emergency was declared on March 13, 2020 (from an average of 502 consults per week, to an average of more than 2000 from April 1). Therefore, we split our sample into two subsamples: before and after March 1, 2020 (24,275 consults before and 12,589 consults after). We obtain similar results (for sensitivity analysis, thus not shown) using February 1, 2020 or March 15 as the cutoff.

### Methods

We compare the distribution of different patient and visit characteristics by calculating their relative share of all consults in each period. While we do not exclude any chief complaint from our main sample, due to space constraints, exhibits only show data for the 15 most common complaints (before and after March 2020). These complaints account for 57.0 percent of all consults until February 2020 and 66.3 percent of cases after March 2020. We calculate standard errors analytically, except for the odds ratio, for which we estimate standard errors using the bootstrap method, stratified by period. Details of the bootstrap method are discussed in the on-line supplementary material.

### Limitations

Our study had a number of limitations. First, our sample is not representative of the US population and therefore results should not be interpreted as such. Second, we are unable to follow patients outside of their interactions with the app and therefore cannot study patient compliance with the recommendations or the resulting patient outcomes. In particular, we cannot speak to the interaction of chat-based consults and care occurring in other settings and therefore cannot assess how cases were managed after the consult or how consults affected the demand for other modes of care.^8,9^

## Results

### Patient and Visit Characteristics

Exhibit 2 shows patient and visit characteristics before and after March 2020. During the baseline period of April 2019 through February 2020, most patients were female (75.1 percent) and between 18–44 years old (88.9 percent of all patients, 90.1 of female and 85.2 of male). Most consults were requested on a weekday (73.5 percent) and during the daytime, defined as 7 a.m.–7 p.m. patient local time (70.8 percent). Since March 2020, there has been a significant increase in the proportion of male patients among users (from 24.9 before to 29.3 percent after), and in patients 45 and older of both genders (from 11.1 percent to 16.9 percent overall; from 14.8 percent to 19.5 percent male; from 9.9 percent to 15.7 percent female). Supplementary Exhibit 1 shows kernel density estimates of the distribution of patient age, by patient gender, before and after March 2020. Since March 2020, there have been slightly more weekday consults (75.4 percent) and daytime consults (73.1 percent).

### Reported Symptoms

Until February 2020, the most common complaints were vaginal discharge, abdominal pain, dysuria, sore throat, cough, headache, anxiety, back pain, depressed mood, fever, chest pain, nasal congestion, dental pain, tender sinuses, nausea, rash, ear pain, and dyspnea. These complaints are largely associated with minor acute conditions, but notably include mental health complaints. Together, these complaints accounted for 57 percent of consults. Panel A of Exhibit 3 shows the individual frequencies of these most common complaints.

The COVID-19 emergency and the period following it were associated with a spike in self-reported symptoms related to COVID-19. Panel B of Exhibit 3 shows the change in symptom frequency from before to after March 2020, sorted by the size of the change. Notable increases were in complaints associated with COVID-19 symptoms: dyspnea, also known as shortness of breath, (OR 4.64, 95 percent CI: 4.11, 5.28) increased from the eighteenth most common complaint to the third most common complaint, and cough (OR 1.98; 95 percent CI: 1.83, 2.16) increased from being the fifth most common complaint to the most common complaint overall. Anxiety (OR 2.00; 95 percent CI: 1.83, 2.20) and depressed mood (OR 1.35; 95 percent CI: 1.20, 1.51) also increased, to become the second and seventh most common complaints. Another set of complaints that saw an increase include those that would typically be addressed elsewhere, such as dental pain (OR 1.41; 95 percent CI: 1.22, 1.62). Some complaints, such as back pain, that were common until February 2020 saw marked decreases from March 2020 (OR 0.55; 95 percent CI: 0.47, 0.64), perhaps due to decreased physical strain due to lockdowns and unemployment.

### Case Resolutions

Most chat-based consults are resolved during the consult by the physician without the patient being further referred by the physician to other providers. Exhibit 4 shows the distribution of physician recommendations. Panel A shows the overall shares of cases that were resolved and referred, before and after March 2020. Before March 2020, a share of 67 percent of cases were resolved. Panels B and C further show the shares of more specific recommendations, separately for resolved and referred cases. The most common resolutions were prescriptions for drugs (34.5 percent of all consults), over-the-counter drugs (18.4 percent), and reassurance (12.9 percent). Lab tests were rarely ordered (1.2 percent). The most common referrals were to primary care physicians (11.9 percent of all consults) and physician specialists (10.8 percent). A small share of cases was referred to emergency departments (6.6 percent) and urgent care (3.7 percent).

Since March 2020, the share of resolved cases increased from 67 to 77.6 percent of consults, with a clear shift toward more prescription drugs (51.2 percent of all consults) and fewer referrals to office-based physicians (6.9 percent to primary care physicians and 7.7 percent to other physicians’ specialists). There was a smaller decrease in the share of consults referred to the emergency room (5.3 percent) and urgent care (2.5 percent).These differences are all statistically significant (p<0.01), based on a two-sided chi-squared test (See supplementary material for more details).

Exhibit 5 shows the distribution of physician recommendations by chief complaint, for the most common complaints, before and after March 2020. After March 2020, there were more case resolutions through prescriptions and fewer referrals across many different complaints, consistent with the circumstances that made referrals to face-to-face encounters less appealing in the presence of stay-at-home orders. Exhibit 5 also reveals the heterogeneity in the resolution and referral rates of different complaints. Complaints associated with minor infections, such as tender sinuses, were more likely than other complaints to be treated remotely via prescriptions both until February 2020 (68.6 percent of patients reporting tender sinuses were prescribed medications, compared with 34.5 of all) and after March 2020 (76 percent of patients reporting tender sinuses were prescribed medications, compared with 51.2 percent of all cases). In contrast, cases with mental health complaints were more likely than other cases to be referred both until February 2020 (62.5 percent of anxiety complaints, compared with 22.7 percent of all cases) and from March 2020 (43.5 percent of anxiety complaints, compared with 14.6 percent of all cases).

## Policy Implications

Telemedicine, which had been expanding prior to the current crisis, is assuming a central role in the response of many healthcare systems to the COVID-19 crisis. Different modes of telemedicine are being rapidly deployed, and existing ones, like the chat-based remote care we have studied, have seen a rapid surge in use. There have been important policy changes to support this shift, such as lifting of requirement for state licensure during the pandemic.

Our analysis highlights several key opportunities for using patient-initiated remote synchronous chat-based consults with physicians. In normal times, chat-based consults offer high rates of resolution and can expedite care for many minor conditions, while further referring complex cases. Beyond the convenience and savings in travel they offer, chat-based consults may provide a point of contact with healthcare providers for people without medical insurance and thus have the potential to mitigate access issues related to insurance coverage.

In an emergency, chat-based consults can quickly adapt to a surge in utilization and provide patients in any location access to primary medical care, even including areas under lockdown conditions that make visiting providers in person more difficult. The increase in complaints related to COVID-19 and the shift toward more resolution and fewer non-urgent referrals of most cases suggest that chat-based remote care has contributed to remote triage in the COVID-19 emergency, internalizing the increased associated risks. The fact that telemedicine clinics can serve patients in all states has significant implications not only for their use during lockdowns but also for handling the surge of deferred non-urgent conditions that will follow once the restrictions are eased. The increase in mental health complaints also suggests that chat-based platforms can also help provide improved access and initial assessment to such cases and may help avert a mental health crisis that might result from extended lockdowns and increased individual economic uncertainty.^10,11^

## Conclusion

Telemedicine serves several purposes in the current emergency, including remote triage and continued care for chronic patients who are particularly vulnerable to COVID-19 complications.^12^ It is likely that care models involving remote care will continue to develop during the current crisis and will assume a greater role than they did previously after the crisis ends.^2,3,13^ These developments raise many questions about the characteristics of different models of telemedicine and their potential role in healthcare provision.

This study provides novel data on the use of chat-based consults with physicians before and during the current public health emergency. Results suggest this type of service is already providing treatment for minor acute cases. While we cannot directly assess provider decision making, the ways physicians have changed their recommendations in response to the constraints of lockdowns (e.g., by prescribing more and referring less to primary care, while still routing some cases to the emergency department) and the variation across conditions in the type of resolution (no referrals of nasal congestion complaints but many referrals of anxiety complaints) provide hopeful signs regarding care quality in the remote setting.

One concern is that the ease of access of direct-to-consumer chat consults could increase utilization by patients with very minor conditions that would otherwise resolve on their own and may even increase low-value use of other services if such cases are unnecessarily referred to other providers. The high rates of remote resolutions and the low rates of referrals (even prior to the COVID-19 crisis) are hopeful signs that this might not be the case in our sample and that providers acknowledge that the distribution of cases arriving through this mode may be different than those arriving at the physician’s office, although further research on this issue is needed.

Given the large expansion of the use of telemedicine, including use by demographic groups of patients (and in other settings, of providers) who were less likely to use such services before the crisis, the current crisis will likely accelerate the adoption of these new modes after the current crisis ends.^13^ Our results highlight the possibility of introducing direct-to-consumer chat-based remote care into the set of care options. Such option improves access and economizes on travel for patients living in remote areas or that are immobile and appears ready to handle minor acute conditions that require little coordination. A bigger challenge, and perhaps a bigger opportunity, is the use of direct-to-consumer remote care to manage chronic and more complex patients. Such patients use care more often and therefore would benefit more from the reduction of travel and the protection from risks (such as COVID-19 and other infections) associated with face-to-face care. However, managing complex cases involves greater coordination challenges and would require tighter integration with other care modes. Direct-to-consumer remote care may also alter the boundaries of local markets for primary care services, by allowing providers to interact with geographically dispersed patients. As this and other new modes of telemedicine are likely going to play an increasing role in the provision of healthcare, its continuing assessment will be an important input for policy and practice alike.

## Data Availability

The data used in this study contain individual administrative data (health insurance and electronic health records) which we obtained from K Health. These data contain confidential patient medical information. Therefore, data cannot be posted online. However, we should be able to share the data with interested researchers for replication related purposes, under the appropriate arrangements for protecting patient confidentiality.

## Acknowledgments

The study was approved by the Ethics Committee of Tel Aviv University (Reference 2020-1440-1). To protect the privacy of patients, we do not report any statistics on groups of fewer than ten patients. Thanks to the sample size, such cells were never encountered in the analysis. Dan Zeltzer acknowledges support from the Foerder Institute for Economic Research at Tel Aviv University.

## Main Exhibits

**Exhibit 1.**
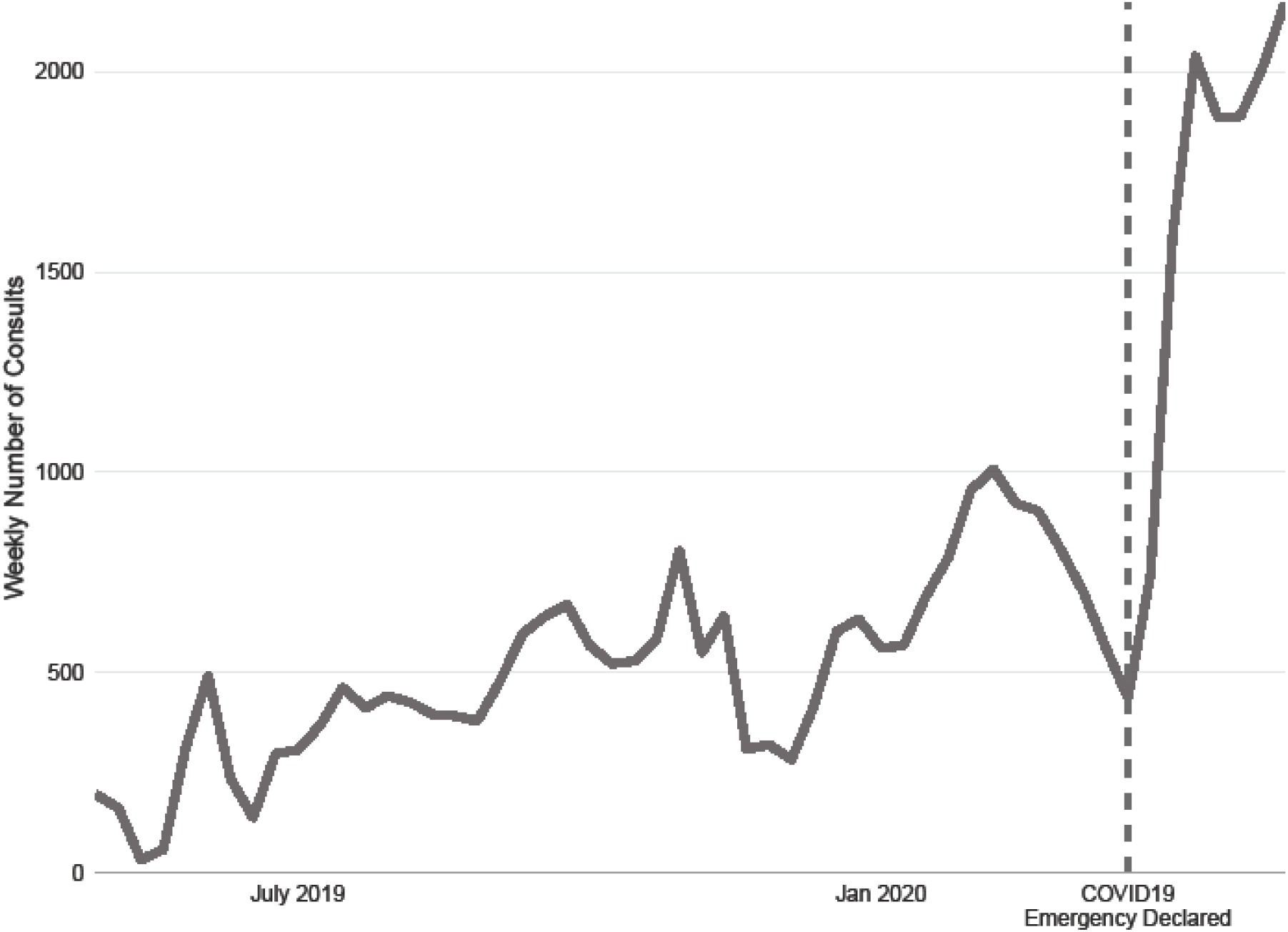
Weekly numbers of completed chat-based consults between April 2019 and April 2020. **Source:** Authors’ analysis of data from K Health electronic health records. **Notes:** Counts refer to completed chat-based remote consults with a physician. See text for detailed sample definition. The dashed line marks the week of March 13, the time when the United States declared COVID-19 to be a national emergency

**Exhibit 2.** Visit and patient characteristics for patient-initiated chat-based consults at the K Health-affiliated clinic from April 2019 through April 2020. **Source:** Authors’ analysis of data from K Health electronic health records. **Notes:** Through 2020 and From March 2020 refer to the periods April 2019–February 2020 (N = 24,275 consults) and March–April 2020 (N = 12,589 consults). Visit time is the time a patient completed the intake and entered the queue to chat with a physician. All differences are significant with *p*-value < 0.01, using two-sided chi-squared test (for details, see supplementary material). Detailed data on the distribution of patient age by gender are available in Supplementary Exhibit 1. Detailed data on consult time are available in Supplementary Exhibit 2.

|  | Through Feb 2020 | From March 2020 |
| --- | --- | --- |
|  | Percent | Percent |
| **A. Patient Characteristics** |  |  |
| Age (years) |  |  |
| 18–44 | 88.9 | 83.1 |
| 45–65 | 11.1 | 16.9 |
| Sex |  |  |
| Female | 75.1 | 70.7 |
| Male | 24.9 | 29.3 |
| **B. Consult Time (patient local time)** |  |  |
| Day of Week |  |  |
| Monday–Friday | 73.5 | 75.6 |
| Saturday–Sunday | 26.5 | 24.6 |
| Time of Day |  |  |
| Day (7 a.m. – 7 p.m.) | 70.8 | 73.1 |
| Night (7 p.m. – 7 a. m.) | 29.2 | 26.9 |

**Exhibit 3.**
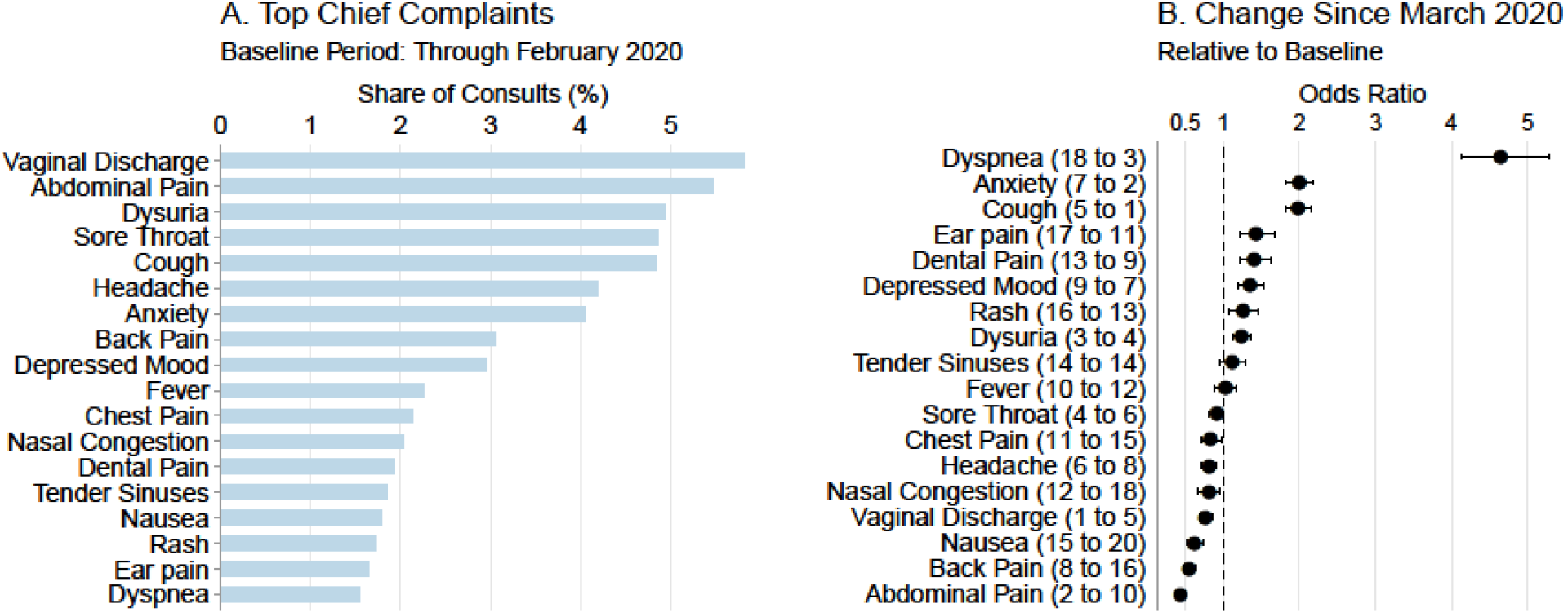
Baseline shares of chief complaints associated with chat consults at the K Health-affiliated clinic and the change in these shares around March 2020. **Source:** Authors’ analysis of data from K Health electronic health records. **Notes:** Panel A shows the frequencies of the most common chief complaints in the baseline period of April 2019 through February 2020, sorted by their share of total consults in descending order. Panel B shows the change in each complaint’s share of total consults after March 2020, relative to the baseline period, sorted by the rate of change in descending order. Points and bars are point estimates and 95 percent confidence intervals for the odds ratios. The dashed line shows an odds-ratio of 1 (no change). The numbers in parentheses show the rank of each condition before and after March 2020. In both panels, data are shown for all patient-reported chief complaints that were among the 15 most common either before or after March 2020. The data are available in Supplementary Exhibit 3.

**Exhibit 4.**
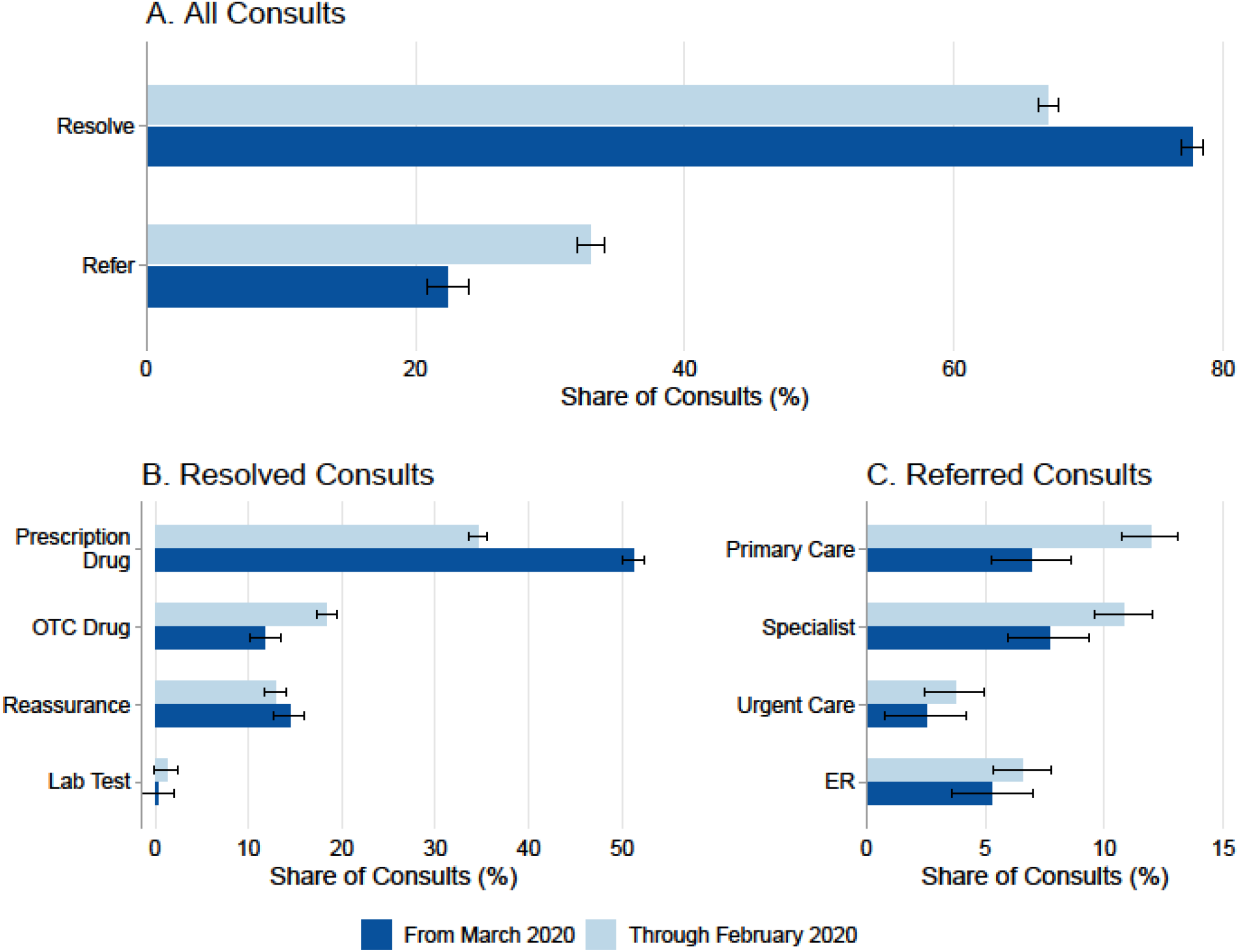
The distribution of resolutions and referrals of chat-based consults with physicians at the K Health-affiliated clinic, before and after March 2020. **Source:** Authors’ analysis of data from K Health electronic health records. **Notes:** For the study sample of completed chat-based remote consults in the K Health-affiliated clinic in the periods of April 2019 through February 2020 and March 2020 through April 2020, Panel A shows the shares of consults for which the physician resolved the case remotely and the share of consults that were further referred to other providers. Panels B and C show further details on the specific type of resolution or referral. In all panels, shares are out of all sampled consults. The data are available in Supplementary Exhibit 4.

**Exhibit 5.**
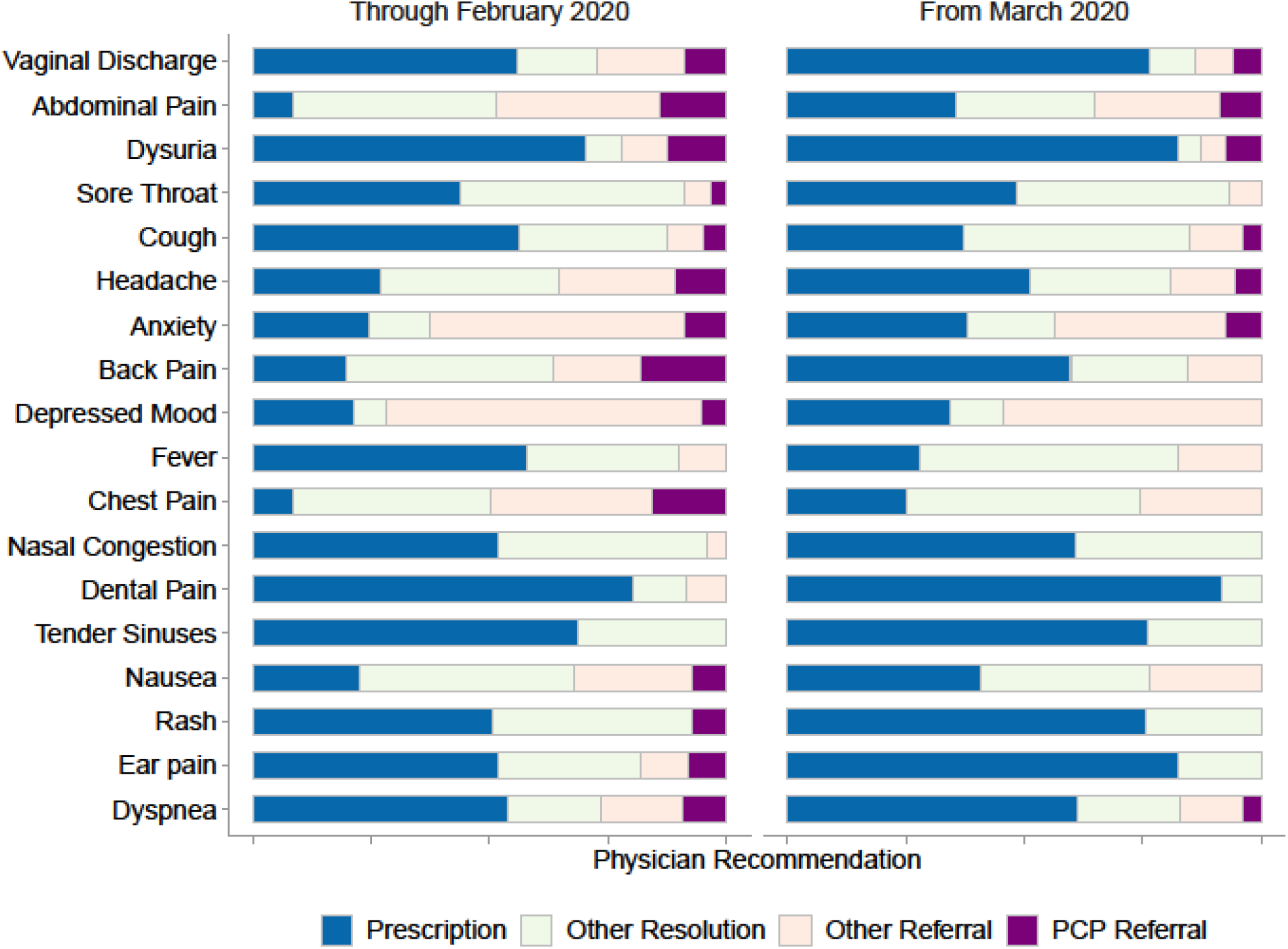
The distribution of resolutions and referrals of chat-based consults with physicians at the K Health-affiliated clinic, in the periods April 2019 through February 2020 and March through April 2020, for each of the most common patient chief complaints. **Source:** Authors’ analysis of data from K Health electronic health records. **Notes:** For the study sample of completed chat-based remote consults in the K Health-affiliated clinic in the periods of April 2019 through February 2020 (left) and March 2020 through April 2020 (right), the figure shows the fraction of each of the four types of physician recommendations: prescription drug (Prescription), over-the-counter drug, reassurance, lab order (Other Resolution), and primary care referral (PCP Referral). The data are available in Supplementary Exhibit 5.

## Supplementary Material

### Details of Statistical Methods

This section describes in the statistical methods we use for estimation and inference. Standard errors of proportions that are presented in our main exhibits were calculated using the formula: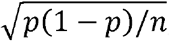, where *n* is the sample size and *p* is the proportion of cases. For comparing proportions (e.g., patient gender) between our two main samples of cases that occurred before and after March 2020, we use a chi-square test for the difference between proportions without continuity correction, with periods (before and after March 2020) as rows, and outcomes (e.g., different age groups or times of consults) as columns. For estimating the standard errors of the odds-ratio of the change in symptom frequencies before versus after March 2020, we use the bootstrap method.^14^ We draw with replacement 1,000 bootstrap samples of the same size as the original data, stratified by period (namely, sampling is done from each of the sampled periods—before and after March 2020—separately). We then calculate, for each drawn sample, the odds ratio. We then report as the confidence interval the 2.5 and 97.5 percentiles of the odds ratio statistic. We implement this method using the “boot” package in R.

## Supplementary Exhibits

**Supplementary Exhibit 1.**
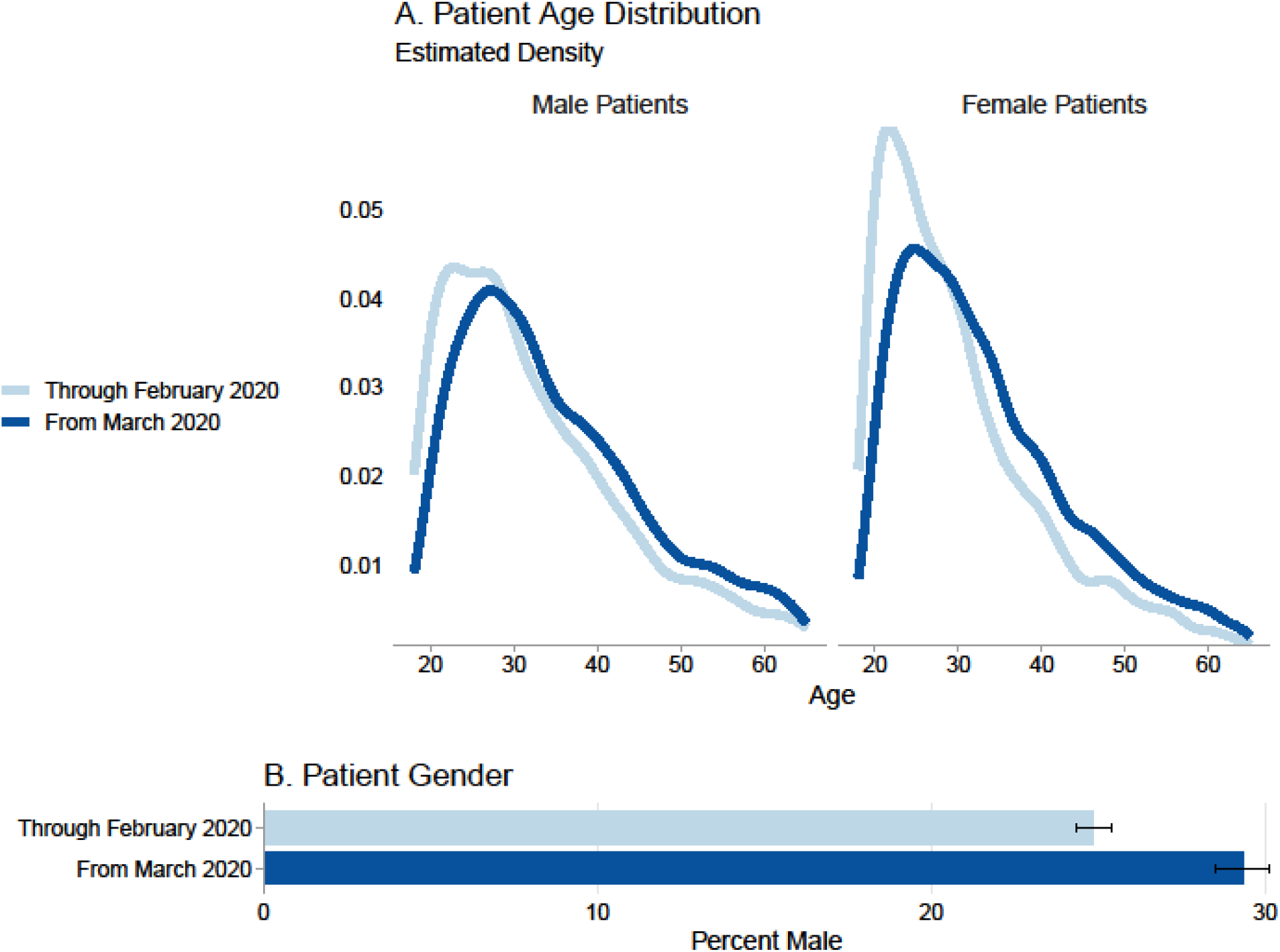
Distribution of patient age, by patient gender, before and after March 2020. **Source:** Authors’ analysis of data from K Health electronic health records. **Notes:** Through Feb 2020 and From March 2020 refer to the periods April 2019 through February 2020 (N = 24,275 consults) and March through April 2020 (N = 12,589 consults).

**Supplementary Exhibit 2.**
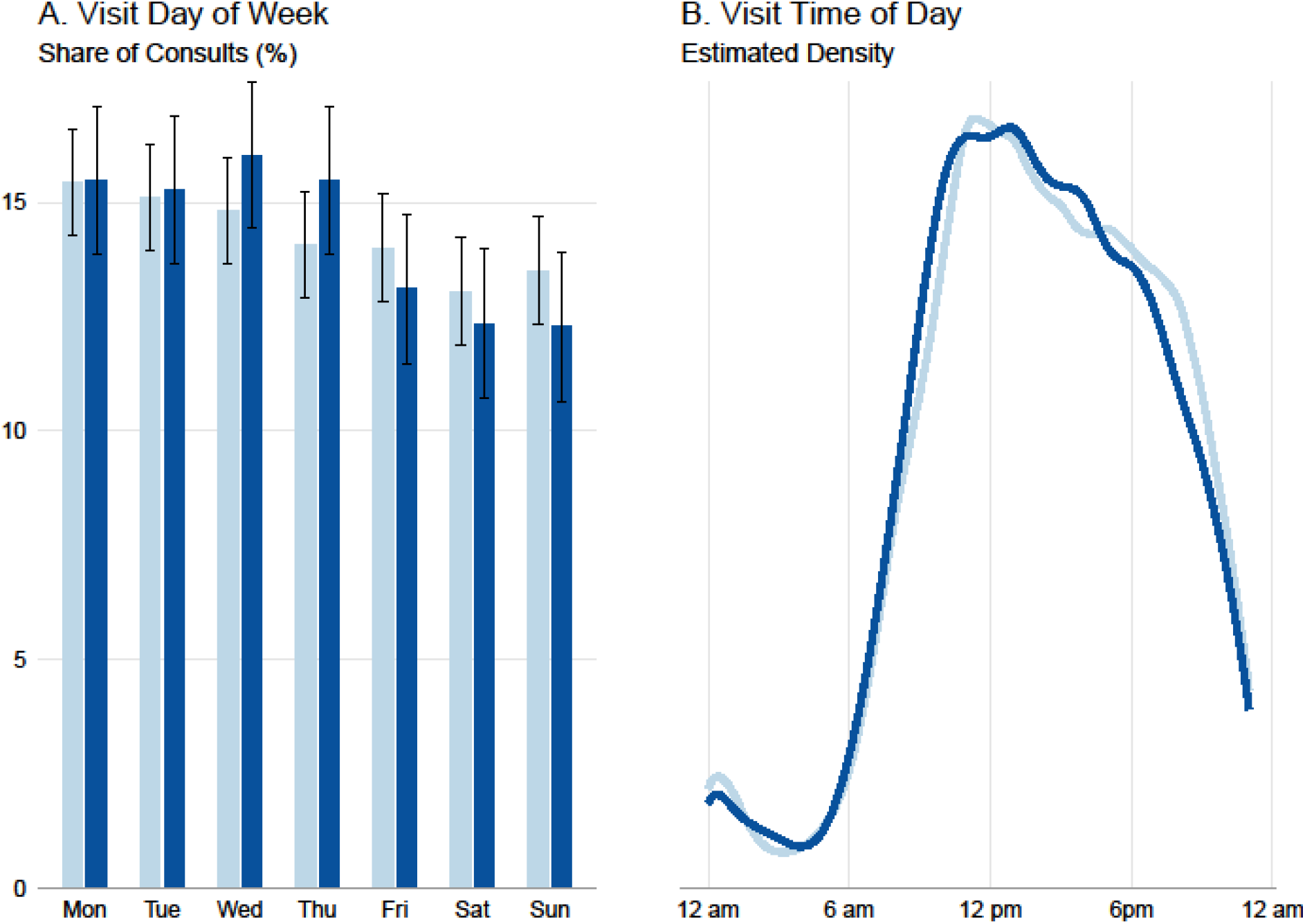
Distribution of visit day of week and time of day, before and after March 2020. **Source:** Authors’ analysis of data from K Health electronic health records. **Notes:** Before March 2020 (light blue lines) and From March 2020 (dark blue lines) refer to the periods April 2019 through February 2020 (N = 24,275 consults) and March through April 2020 (N = 12,589 consults).

**Supplementary Exhibit 3.**
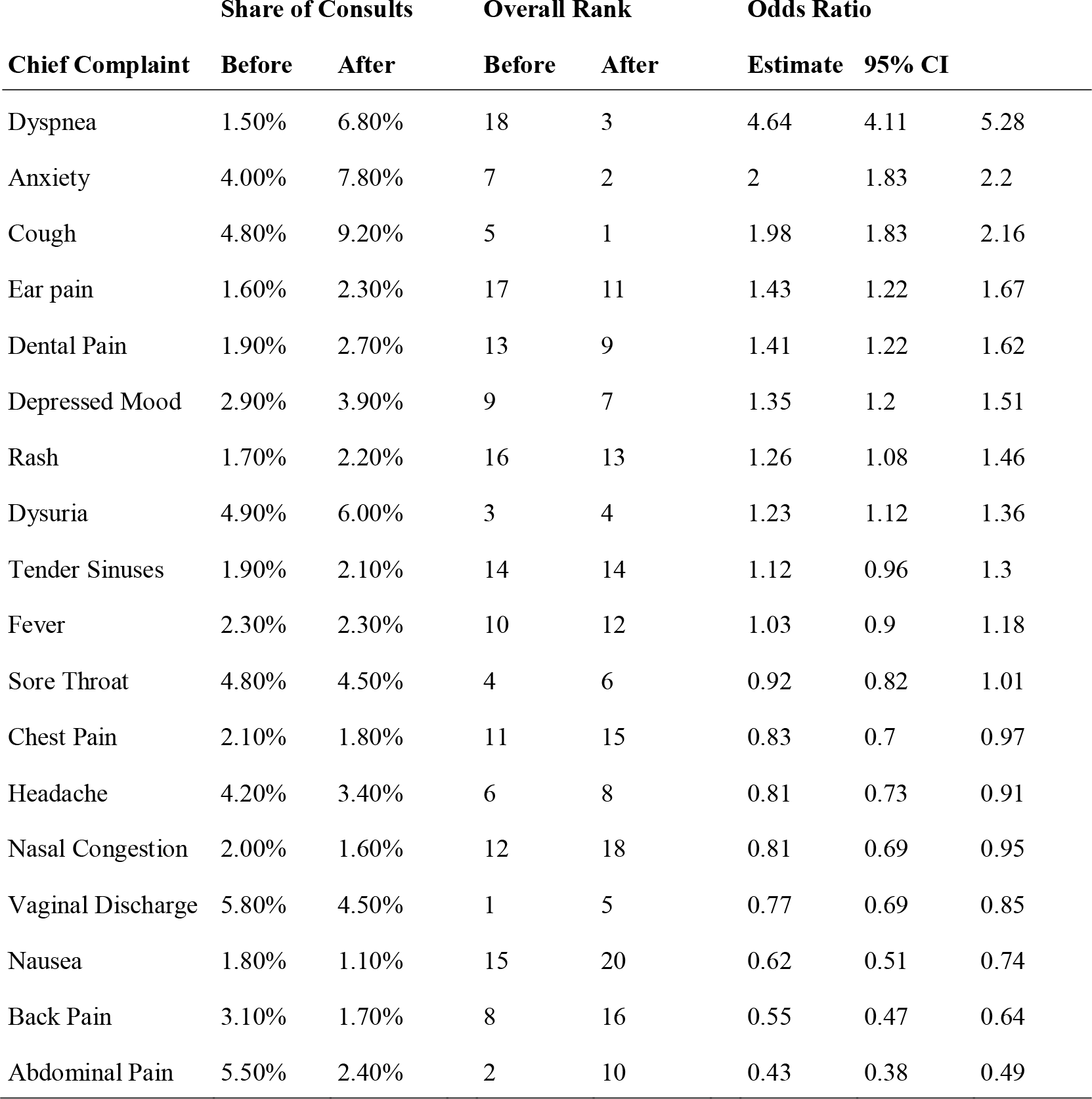
Frequency of the 15 most common chief complaints, before and after March 2020. **Source:** Authors’ analysis of data from K Health electronic health records. **Notes:** The table shows the share, out of all consults, of each of the most common chief complaints in the periods of April 2019 through February 2020 and March through April 2020. Complaints are sorted by their share of total consults in the baseline period in descending order. Rank is the frequency rank (with 1 being the most frequent). Odds ratios reflect the change in the share of each complaint before and after March 2020. Data are shown for all patient-reported chief complaints that were among the 15 most common either before or after March 2020.

**Supplementary Exhibit 4.**
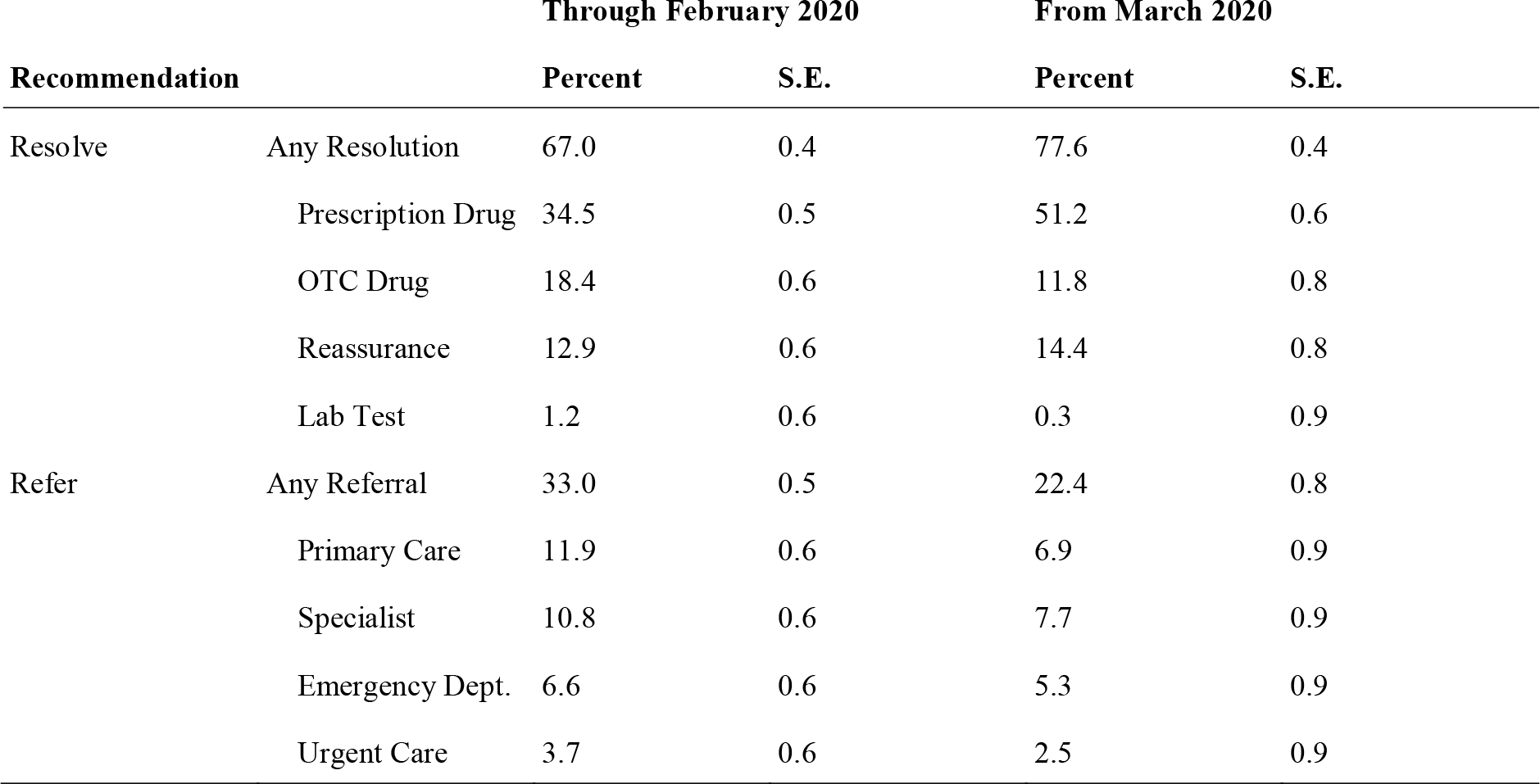
Distribution of Physician Recommendations (for All Cases), Before and After March 2020. **Source:** Authors’ analysis of data from K Health electronic health records. **Notes:** For the study sample of completed chat-based remote consults in the K Health-affiliated clinic in the periods of April 2019 through February 2020 and March 2020 through April 2020, the table shows the shares of consults with each specific type of physician resolution or referral. OTC drugs refers to over-the-counter drugs. Any resolution is the sum of all types of resolutions; any referral is the sum of all types of referrals.

**Supplementary Exhibit 5.** Physician recommendations stratified by complaint, for the 15 most common chief complaints before and after March 2020. **Source:** Authors’ analysis of data from K Health electronic health records. **Notes:** For the study sample of completed chat-based remote consults in the K Health-affiliated clinic in the periods of April 2019 through February 2020 (left) and March 2020 through April 2020 (right), the table shows the fraction of each of four types of physician recommendations: prescription drug (Prescription), over-the-counter drug, reassurance, lab order (Other Resolution), and primary care referral (PCP Referral).

| Chief Complaint | Through February 2020 |  |  |  |  | From March 2020 |  |  |  |
| --- | --- | --- | --- | --- | --- | --- | --- | --- | --- |
|  | Rank | Prescription | Other Resolution | Other Referral | PCP Referral | Prescription | Other Resolution | Other Referral | PCP Referral |
| Vaginal Discharge | 1 | 55.8 | 16.9 | 18.4 | 8.9 | 76.5 | 9.4 | 8.1 | 6 |
| Abdominal Pain | 2 | 8.5 | 43.1 | 34.5 | 14 | 35.6 | 29.1 | 26.5 | 8.8 |
| Dysuria | 3 | 70.1 | 7.8 | 9.4 | 12.7 | 82.3 | 4.8 | 5.5 | 7.4 |
| Sore Throat | 4 | 43.7 | 47.4 | 5.6 | 3.2 | 48.3 | 45 | 6.7 |  |
| Cough | 5 | 56.2 | 31.2 | 7.8 | 4.8 | 37.3 | 47.6 | 11.4 | 3.8 |
| Headache | 6 | 27.1 | 37.6 | 24.3 | 11 | 51.3 | 29.4 | 13.8 | 5.6 |
| Anxiety | 7 | 24.6 | 12.9 | 53.6 | 8.9 | 38.1 | 18.4 | 35.9 | 7.5 |
| Back Pain | 8 | 19.6 | 43.8 | 18.4 | 18.2 | 59.8 | 24.6 | 15.6 |  |
| Depressed Mood | 9 | 21.2 | 7 | 66.2 | 5.5 | 34.4 | 11.3 | 54.3 |  |
| Fever | 10 | 57.9 | 32.1 | 10 |  | 28 | 54.3 | 17.7 |  |
| Chest Pain | 11 | 8.5 | 41.6 | 34.2 | 15.7 | 25.1 | 49.3 | 25.6 |  |
| Nasal Congestion | 12 | 51.8 | 44.1 | 4.2 |  | 60.7 | 39.3 |  |  |
| Dental Pain | 13 | 80.4 | 10.9 | 8.7 |  | 91.6 | 8.4 |  |  |
| Tender Sinuses | 14 | 68.6 | 31.4 |  |  | 76 | 24 |  |  |
| Nausea | 15 | 22.6 | 45.3 | 24.7 | 7.4 | 40.9 | 35.6 | 23.5 |  |
| Rash | 16 | 50.8 | 41.8 |  | 7.5 | 75.8 | 24.2 |  |  |
| Ear pain | 17 | 51.9 | 30.1 | 9.9 | 8.1 | 82.4 | 17.6 |  |  |
| Dyspnea | 18 | 53.7 | 19.8 | 17.1 | 9.4 | 61.3 | 21.4 | 13.2 | 4.1 |

